# A collaborative online AI engine for CT-based COVID-19 diagnosis

**DOI:** 10.1101/2020.05.10.20096073

**Authors:** Yongchao Xu, Liya Ma, Fan Yang, Yanyan Chen, Ke Ma, Jiehua Yang, Xian Yang, Yaobing Chen, Chang Shu, Ziwei Fan, Jiefeng Gan, Xinyu Zou, Renhao Huang, Changzheng Zhang, Xiaowu Liu, Dandan Tu, Chuou Xu, Wenqing Zhang, Dehua Yang, Ming-Wei Wang, Xi Wang, Xiaoliang Xie, Hongxiang Leng, Nagaraj Holalkere, Neil J. Halin, Ihab Roushdy Kamel, Jia Wu, Xuehua Peng, Xiang Wang, Jianbo Shao, Pattanasak Mongkolwat, Jianjun Zhang, Daniel L. Rubin, Guoping Wang, Chuangsheng Zheng, Zhen Li, Xiang Bai, Tian Xia

## Abstract

Artificial intelligence can potentially provide a substantial role in streamlining chest computed tomography (CT) diagnosis of COVID-19 patients. However, several critical hurdles have impeded the development of robust AI model, which include deficiency, isolation, and heterogeneity of CT data generated from diverse institutions. These bring about lack of generalization of AI model and therefore prevent it from applications in clinical practices. To overcome this, we proposed a federated learning-based Unified CT-COVID AI Diagnostic Initiative (UCADI, http://www.ai-ct-covid.team/), a decentralized architecture where the AI model is distributed to and executed at each host institution with the data sources or client ends for training and inferencing without sharing individual patient data. Specifically, we firstly developed an initial AI CT model based on data collected from three Tongji hospitals in Wuhan. After model evaluation, we found that the initial model can identify COVID from Tongji CT test data at near radiologist-level (97.5% sensitivity) but performed worse when it was tested on COVID cases from Wuhan Union Hospital (72% sensitivity), indicating a lack of model generalization. Next, we used the publicly available UCADI framework to build a federated model which integrated COVID CT cases from the Tongji hospitals and Wuhan Union hospital (WU) without transferring the WU data. The federated model not only performed similarly on Tongji test data but improved the detection sensitivity (98%) on WU test cases. The UCADI framework will allow participants worldwide to use and contribute to the model, to deliver a real-world, globally built and validated clinic CT-COVID AI tool. This effort directly supports the United Nations Sustainable Development Goals’ number 3, Good Health and Well-Being, and allows sharing and transferring of knowledge to fight this devastating disease around the world.

## Introduction

COVID-19 has become a global pandemic. RT-PCR was adopted as the main diagnostic modality to detect viral nucleotide in specimens from patients with suspected COVID-19 infection and remained as the gold standard for active disease confirmation. However, due to the greatly variable disease course in different patients, the detection sensitivity is only 60%-71% ^1-3^ leading to considerable false negative results. These symptomatic COVID 19 patients and asymptomatic carriers with false negative RT-PCR results pose a significant public threat to the community as they may be contagious. As such, clinicians and researchers have made tremendous efforts searching for alternative and/or complementary modalities to improve the diagnostic accuracy for COVID-19.

COVID-19 patients present with certain unique radiological features on chest computed tomography (CT) scans including ground glass opacity, interlobular septal thickening, consolidation etc., that have been used to differentiate COVID-19 from other bacterial or viral pneumonia or healthy individuals^4-7^. CT has been utilized for diagnosis of COVID-19 in some countries and regions with reportedly sensitivity of 56-98%^2,3^. However, these radiologic features are not specifically tied to COVID-19 pneumonia and the diagnostic accuracy heavily depending on radiologists’ experience. Particularly, insufficient empirical understanding of the radiological morphology characteristic of this unknown pneumonia resulted in inconsistent sensitivity and specificity by varying radiologists in identifying and assessing COVID-19. A recent study has reported substantial differences in the specificity in differentiation of COVID-19 from other viral pneumonia by different radiologists^8^. Meanwhile, CT-based diagnostic approaches have led to substantial challenges as many suspected cases will eventually need laboratory confirmation. Therefore, there is an imperative demand for an accurate and specific intelligent automatic method to help to address the clinical deficiency in current CT approaches.

Successful development of an automatic method depends on a tremendous amount of imaging data with high quality clinical annotation for training an artificial intelligence (AI) model. We confronted several challenges for developing a robust and universal AI tool for precise COVID-19 diagnosis: 1) data deficiency. Our high-quality CT data sets were only a small sampling of the full infected cohorts and therefore it is unlikely we captured the full set radiological features. 2) data isolation, Data derived across multiple centers was difficult to transfer for training due to security, privacy, and data size concerns. and 3) data heterogeneity. Datasets were generated by different scanner machines which introduces an additional layer of complexity to the training because every vendor provides some unique capabilities. Furthermore, it is unknown whether COVID-19 patients in diverse geographic locations, ethnic groups, or demographics show similar or distinct CT image patterns. All of these may contribute to a lack of generalization for an AI model, which a serious issue for a global AI clinical solution.

To solve this problem, we propose here a Unified CT-COVID AI Diagnostic Initiative (UCADI) to deliver an AI-based CT diagnostic tool. We base our developmental philosophy on the concept of federated learning, which enables machine learning engineers and medical data scientists to work seamlessly and collectively with decentralized CT data without sharing individual patient data, and therefore every participating institution can contribute to AI training results of CT-COVID studies to a continuously-evolved and improved central AI model and help to provide people worldwide an effective AI model for precise CT-COVID diagnosis (Fig. 1).

**Figure 1.**
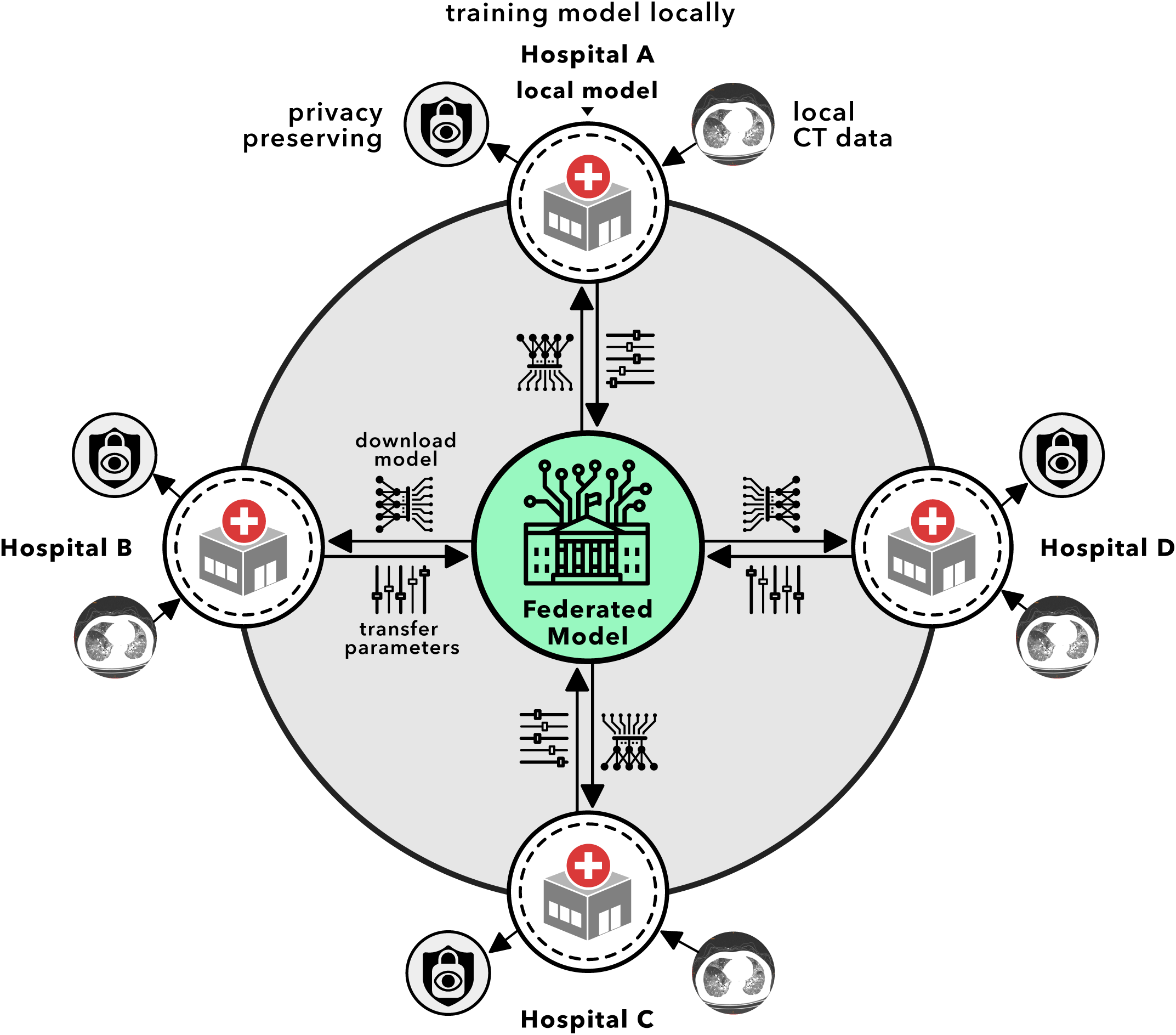
The conceptual architecture of UCADI on the basis of federated learning. UCADI stakeholders firstly download the code and train a new model locally based on the initial model, and secondly transfer the encrypted model parameters back to the federated model. The central server combines the contributions shared from all of the UCADI participants.

**Figure 2.**
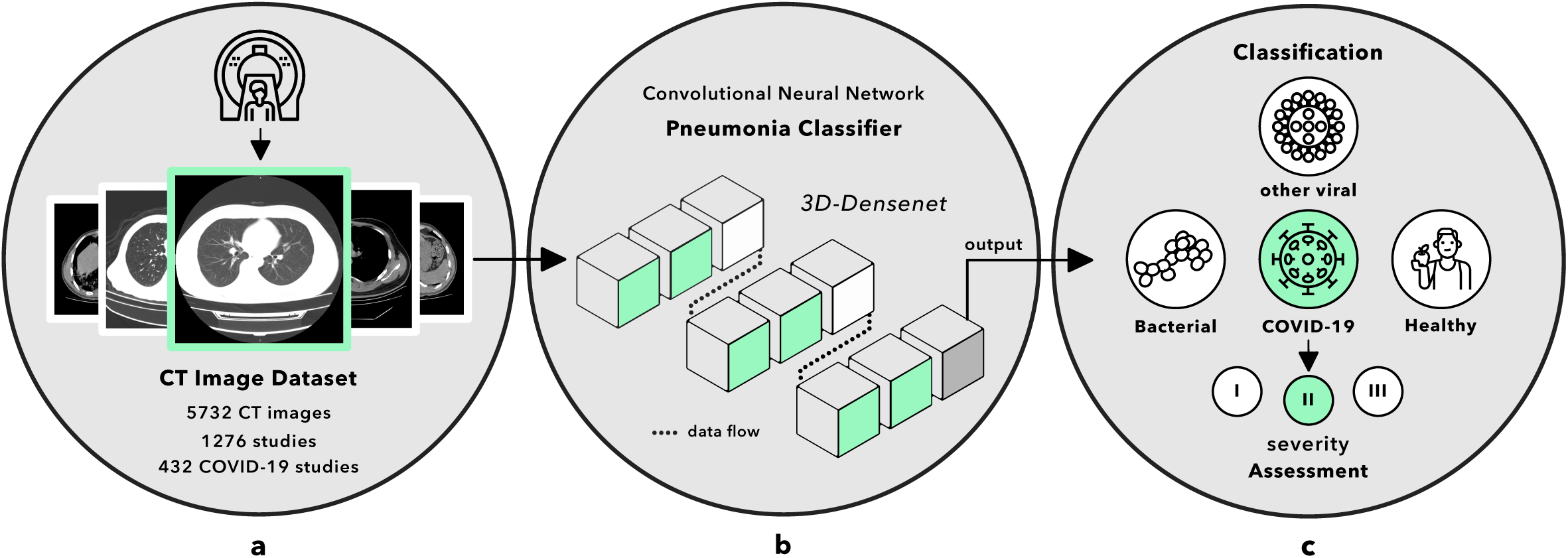
Data and strategy. **a**, number of CT studies and total images. **b**, the CNN was developed based on 3D-Densenet, consisting of 6 dense blocks in green, 2 transmit blocks in white and an output layer in gray. Pre-processed 128-x-128-pixel CT images of one case were fed to the network across 14 3D-convolution layers and a number of functions embedded in 3D blocks, finally received the predicted classification result. **c**, the CNN classified CT case into 4 types and further assessed the severity into I or II or III if the case was predicted as COVID-19.

## Results

### Building AI model using pooled data

We firstly gathered a dataset of 5732 CT images from 1276 individuals collected from multiple centers of Tongji Hospital including Tongji Hospital Main Campus (3457 CT images from 800 studies), Tongji Optical Valley Hospital (882 CT images from 227 studies), and Tongji Sino-French New City Hospital (1393 CT images from 241 studies) (Table 1 for patient information). Among these patients, 432 patients had COVID-19 pneumonia confirmed by RT-PCR; 76 patients had other viral pneumonia including 7 cases with respiratory syncytial virus (RSV), 13 with EB virus, 16 with cytomegalovirus, 3 with influenza A, 1 with parainfluenza virus and 36 with mixed virus pneumonia that were confirmed PCR or antibodies against corresponding viruses; 350 patients had bacterial pneumonia confirmed CT scan and bacterial culture. The remaining 418 individuals having clinical symptoms of respiratory system were healthy individuals who had normal chest CT scans. Based on the dataset, we developed an initial deep learning model by using convolutional neural networks (CNN) (detailed in Methods).

**Table 1.** Patient information of 1276 studies collected from Tongji Hospital regarding gender and age distribution.

|  | Male | Female | 0-20<br>years | 20-40<br>years | 40-60<br>years | 60-80<br>years | >80<br>years |
| --- | --- | --- | --- | --- | --- | --- | --- |
| **Patient Number** | 617 | 659 | 40 | 444 | 421 | 340 | 31 |

Next, we validated the predictive performance of the CNN through a classification task: four-class pneumonia partition—four featured clinical diagnoses in determining suspected cases of COVID-19. This task aimed at distinguishing COVID-19 (Fig. 3. i) from three types of non-COVID-19 (Fig. 3. ii) including other viral pneumonia, bacterial pneumonia, and healthy cases (d, e, and f in Fig. 3). We selected 20% of 1036 CT cases in training and validation set for 5-fold cross-validation. The CNN demonstrated the validation result that achieved overall sensitivity of 77.2% and specificity of 91.9%.

**Figure 3.**
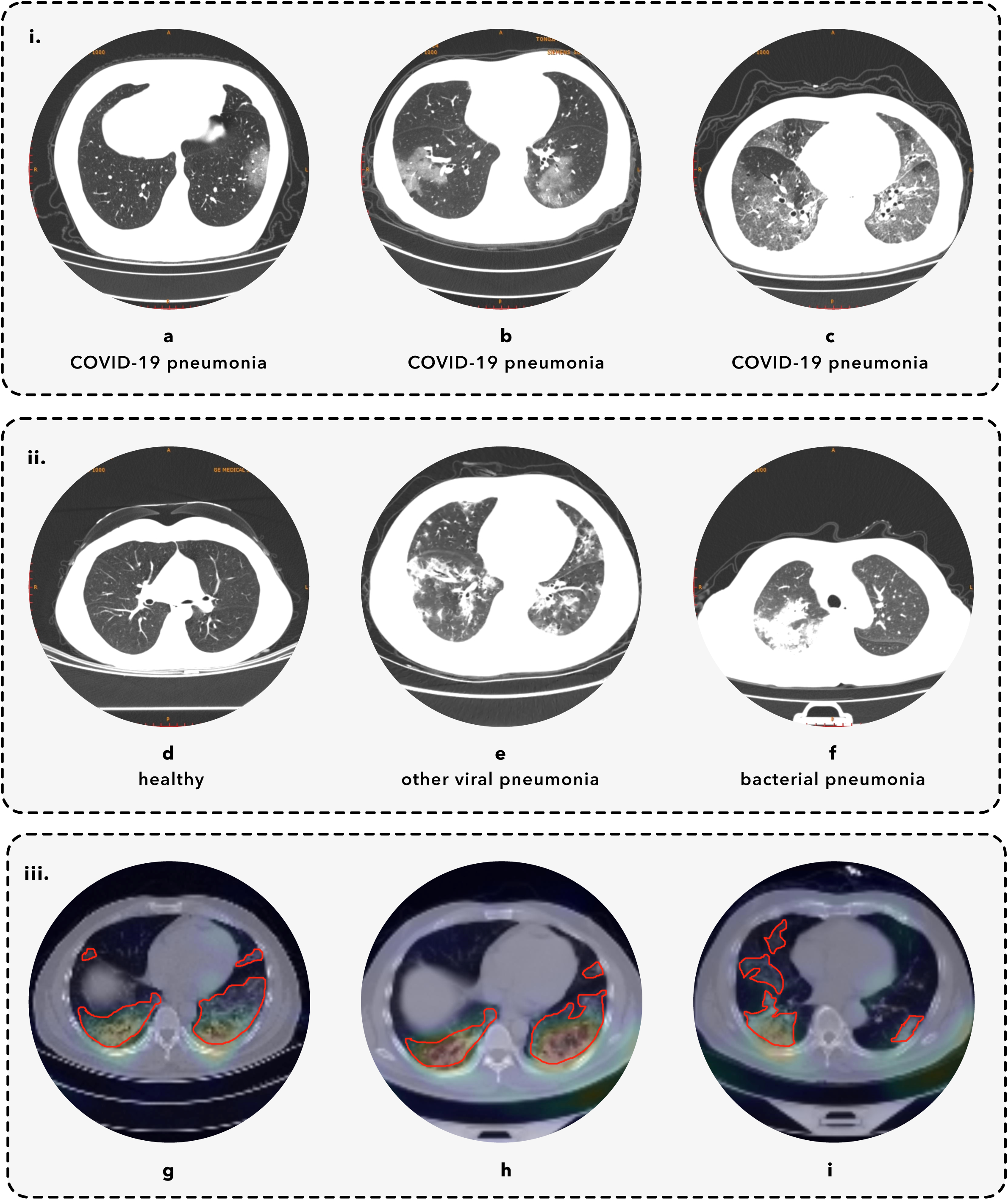
CT images. i and ii, the taxonomy of pneumonia and featured CT image for per-class. iii, the heatmap generated by GradCAM and local lesions annotated by the radiologist. **i**, COVID-19 pneumonia. **a**, **b**, **c** represent the CT images of COVID-19 defined by radiological features. ii, non-COVID-19 cases. **d**, **e**, **f** respectively displays the CT image of healthy case, other viral pneumonia, and bacterial pneumonia. **iii**, CAM visualized the image areas which lead to classification decision. The radiologist, LYM [9 years’ experience], from Department of Radiology, Tongji Hospital circumscribed the local lesions with the red curved masks. **g**-**h**, patients with COVID-19 pneumonia.

We further tested the previously trained CNN by conducting a comparative study of same task between the CNN and expert radiologists using previously separated test set (detailed in Methods). Six qualified radiologists (ZL [18 years’ experience], LYM [9 years’ experience], YZL [9 years’ experience], COX [8 years’ experience], HLM [4 years’ experience], GC [4 years’ experience]) from department of radiology, Tongji Hospital (Main campus), Wuhan, China were asked to make diagnosis as one of above 4 classes based on CT study. In this task, the CNN achieved a sensitivity of 97.5% and specificity of 89.4% in differentiating COVID-19 from three types of non-COVID-19 cases whereas six radiologists obtained the average 79% in sensitivity (87.5%, 90%, 55%, 80%, 68%, 93%, respectively, and 90% for the maximal voting value among six radiologists), and 90% in specificity (92%, 97%, 89%, 95%, 88%, 79%, respectively, and 95.6% for the maximal voting value) (Fig 4). In the Tongji dataset, the CNN shows performance approaching that of expert radiologists. To examine the reliability of the model, we performed class activation mapping (CAM) analysis for raw CT images in both validation and test datasets^9^ and visualized the featured image regions which lead to classification decision. As shown in Figure 3. iii, the heatmap generated by CAM mostly characterized local lesions suggesting the model learned radiologic features rather than simply overfitting the dataset.

**Figure 4.**
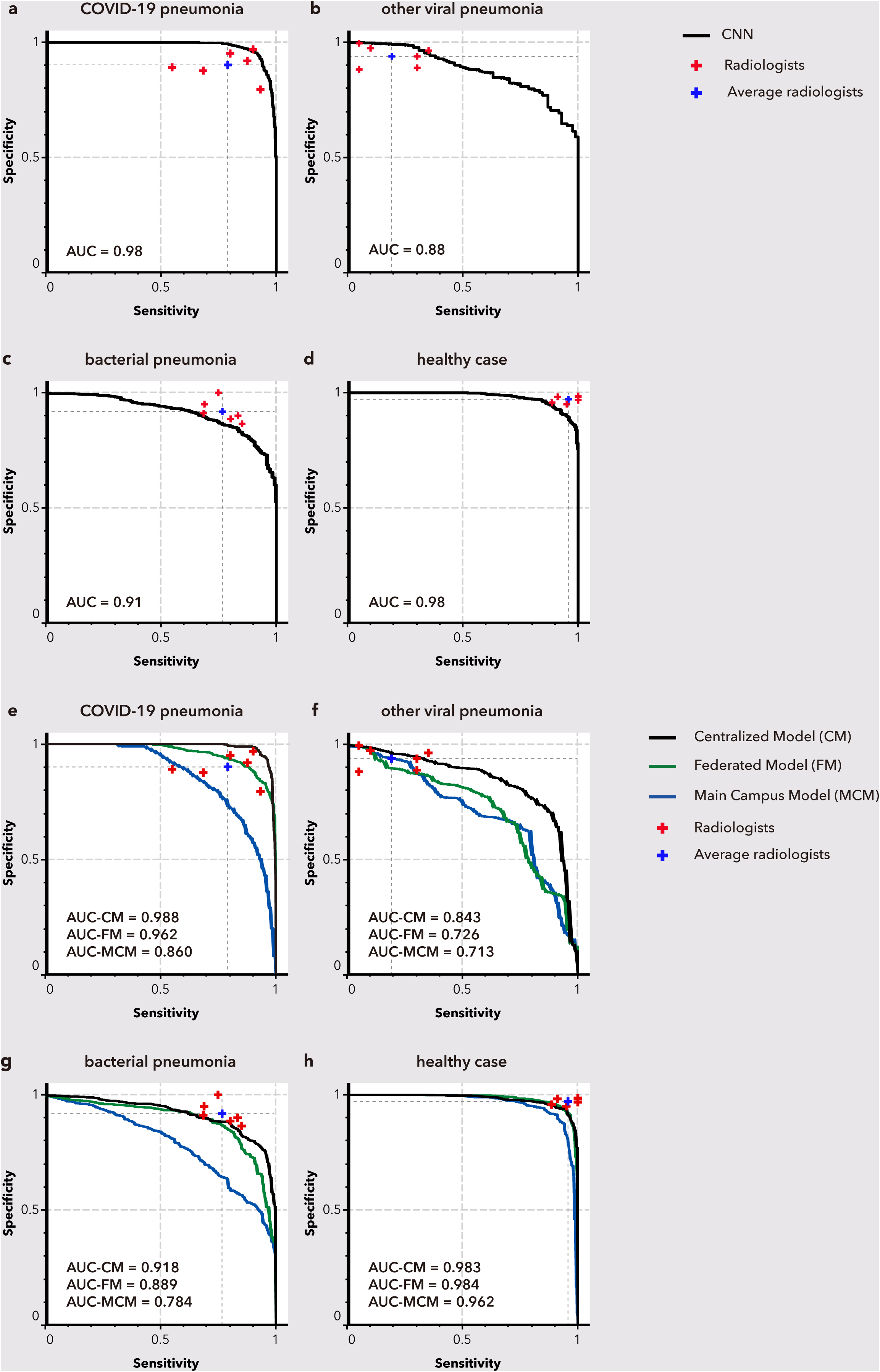
Pneumonia classification performance of CNN models and radiologists. This figure illustrates the comparative analysis between the CNN and radiologists by correlating the ROC curve of CNN and sensitivity-specificity points of six invited radiologists for two conducted classification test tasks. **a-d**, per-class evaluation for three types of pneumonia and healthy case. The curve in black represents the performance of the CNN. Cross marks in red separately represent the performance of six radiologists and the blue mark annotates the average capability, **e-h**, comparative evaluation of centralized-trained initial model, federated model, and Tongji Main Campus model on four per-class classification tasks.

To comprehensively evaluate the comparisons of two tasks, we visualized the correlation of sensitivity and specificity via receiver operating characteristic (ROC) curve to calculate the area under the curve (AUC) for representing the CNN’s classification performance. As a result, the AUC of the CNN attained 0.98, 0.88, 0.91, 0.98 in specifically identifying COVID-19 pneumonia, other viral pneumonia, bacterial pneumonia, and healthy tissue from 4 classes, and 0.92, 0.92, 0.95 in assessing three ordinal severities of COVID-19. Fig. 4 illustrates the ROC curve of the CNN and sensitivity-specificity points displaying radiologists’ diagnosis. Importantly, the CNN performed comparable sensitivity-specificity to all six radiologists in differentiating COVID-19 from non-COVID-19 cases (Fig. 4a). Meanwhile, the CNN also performed equivalent sensitivity-specificity in comparison with average radiologists in the assessment of three severities (e, f, g in Fig. 4). However, the CNN revealed insufficient capability in determining other viral pneumonia (Fig. 4b), bacterial pneumonia (Fig. 4c), and healthy case (Fig. 4d).

To test the generalization of the initial model that was trained exclusively on data from Tongji hospitals, we evaluated the predictive performance using CT data from 100 confirmed COVID-19 cases generated at Wuhan Union hospital. The accuracy of the model was only 72%, compared with a 97% sensitivity using reserved testing data from Tongji hospitals. This demonstrated a lack of generalization for the initial model.

### The global online AI diagnostic engine enabled with federated learning

To overcome the hurdle, we proposed a federated learning framework to facilitate UCADI, a global joint effort to generate an AI based on large scale date and integration of diverse ethnic patient groups. In the traditional AI approach, sensitive user data from different sources are gathered and transferred to a central hub where models are trained and generated. The federated learning proposed by Google^10^, in contrast, is a decentralized architecture where the AI model is distributed to and executed at each host institution with the data sources or client ends for training and inferencing. The local copies of the AI model on the host institution eliminate network latencies and costs incurred due to sharing large size of data with the central server. Most importantly, the strategy privacy preserved by design enables medical centers collaborating on the development of models, but without need of directly sharing sensitive clinical data with each other.

We implemented the federated learning framework at http://www.ai-ct-covid.team/ where we deployed the initial model to provide 1) online diagnostic interface allowing people easily query the model with patient CT images and 2) AI development federated learning interface(detailed in Methods). UCADI stakeholders can download the code and train a new model based on the initial model. Once the new model had been trained locally for several iterations, if UCADI participants share their updated version of the model, the framework will encrypt the model parameters based on Learning with Errors (LWE)-based encryption^11^ and transfer them back to the centralized server via a customized server protocol. Participants’ datasets will keep within their own secure infrastructure. The central server would then combine the contributions from all of the UCADI participants. The updated model parameters would then be shared with all participants, which enables continuation of local training. The framework is highly flexible, allowing hospitals join or leave the UCADI initiative at any moments, because it is not tied to any specific data cohorts.

With the framework, we deployed two experiments to validate federated learning concept on the CT COVID data. Firstly, we trained three models for each of three Tongji hospital datasets, and then transferred the datasets to three physically independent computer servers, respectively, and trained a Tongji federated model in a simulation mode (detailed in Methods). As shown in Figure 4. e-h, the federated model performed close to the centralized-trained initial model and better than Tongji Main Campus model for predicting COVID-19, bacterial pneumonia and healthy case (the comparison not applied to models of Tongji Sino-French Hospital and Tongji Optics Valley because they lack of other viral pneumonia data). It shows the effectiveness of federated model. In the second experiment, we trained a federated model in real mode based on three Tongji hospital datasets (432 COVID-19 cases) and 407 confirmed COVID-19 cases from Wuhan Union hospital. We tested the federated model performance on predicting the same 100 confirmed Wuhan Union COVID-19 cases which we used to test the initial model previously. The result, 98% sensitivity, was improved compared to the initial model (72% sensitivity) which was centralized trained only based on data from three Tongji hospitals.

## Discussion

COVID-19 is a global pandemic. Over 2 million people have been infected, tens of thousands hospitalized, and nearly 200,000 have died worldwide as of April 23^rd^, 2020. There are borders between countries. But only real border in this war is the border between human being and virus. We need a global joint effort to fight the virus. The first challenge we have confronted in this war is to deliver is deliver people precise and effective diagnosis. In this study, we introduce a globally collaborative AI initiative framework, UCADI, to assist radiologists, streamline, and accelerate CT-based diagnosis. Firstly, we developed an initial CNN model that achieved a performance comparable to expert radiologist in classifying pneumonia to identify COVID-19, and additionally assessing the severity of identified COVID-19. Furthermore, we developed a federated learning framework, based on which hospitals worldwide can join UCADI to jointly train an AI-CT model for COVID-19 diagnosis. With CT data from multiple Wuhan hospitals, we confirmed the effectiveness of this the federated learning approach. We have shared the initial model and the federated learning programmatic API source code (https://github.com/HUST-EIC-AI-LAB/) and encourage hospitals worldwide join UCADI to form an international collaboration to fight the virus with a globally trained AI application. It is worth noting that there is still need for improvement in the technical implementation in the framework: 1) The number of local training iterations before global parameter updating. The number of local training iterations has a direct influence on the training efficiency, effectiveness, and model performance. Currently, different clients in UCADI framework train with their private data for one epoch before sending the parameter gradients to the global server. We will construct more detailed experiments about this hyper-parameter to explore the best trade-off between model performance and communication cost. 2) Private information leakage from gradients. Reconstruction of input data from the parameter gradients is possible for realistic deep architectures, and an encryption-decryption module is needed in the federated learning framework. We have adopted an additively homomorphic encryption scheme in our COVID diagnosis framework. The parameter gradients sent to the global server are encrypted while the secret key is kept confidential from the global server, which guarantees the privacy security of our framework. 3) Non-IID and unbalanced data distribution. The training data available is typically based on the patients in the hospital, and any particular hospital’s local dataset will not be representative of the entire distribution. Therefore, it requires a dynamic aggregation method that aggregates different parameter gradients via dynamic weighted averaging. Hence, it can decrease the influence of non-IID and unbalanced data.

## Methods

### CT data collecting and processing

This study was approved by the Ethics Committee Tongji Hospital, Tongji Medical College of Huazhong University of Science and Technology to access this dataset for research purpose.

Here we list the three major scanners used to obtain CT scans: GE Medical System/LightSpeed16, SOMATOM Definition AS+, and GE Medical Systems/Discovery 750 HD. The scanning protocols of slice thicknesses and reconstruction kernel were 1.25mm and adaptive statistical iterative reconstruction (60%) for two GE scanners whilst 1mm and sinogram affirmed iterative reconstruction for the Siemens scanner. The high-quality CT image data from the 432 COVID-19 patients were scanned, enrolled, selected and annotated in this study since January 7, 2020 while other image data were retrospectively collected from CT databases of the three Tongji Hospitals. In addition, we collected an independent cohort including 507 COVID-19 pneumonia CT cases confirmed by chest CT from Union Hospital, Wuhan, China. The cohort was used for testing the performance of initial model and the multi-hospital model using federated learning framework.

We conducted image processing of the raw CT image data to reduce computing burdens. We utilized a sampling method to select 5 subsets of CT slices from all sequential images of one CT case using random starting positions and scalable sampling intervals on transverse view to picture the infected lung regions. All 5 processed subsets were separately fed to the CNN to obtain average predictive probabilities, which can effectively include impacts of different levels of lung from all CT slices. To further improve computing efficiency, we resized each slice from 512 to 128 pixel regarding its width and height and rescaled the lung windows of CT to a range from -1200 to 600 and normalized them via the Z-score means before feeding the CNN.

### Building AI model using pooled data

The dataset was split out into the training and validation set with 1036 cases (80% for training, 20% for validation), and independent test set with 240 cases consisting of 80 COVID-19 studies (28 from Main Campus Hospital, 30 Sino-French New City Hospital, 20 Optical Valley Hospital), 20 with other viral pneumonia (19 from Main Campus Hospital, 1 Sino-French New City Hospital), 60 with bacterial pneumonia (50 from Main Campus Hospital, 8 Sino-French New City Hospital, 2 Optical Valley Hospital), and 80 healthy cases (58 Main Campus Hospital, 10 Sino-French New City Hospital, 12 Optical Valley Hospital). We particularly considered the balanced data distribution of 4 classes in test set. We initially trained a four-class CNN (Fig. 2) based on 3D-Densenet^12^, a densely connected convolutional network, which performed remarkable advantages in classifying CT images. We customized its architecture to contain 14 3D-convolution layers distributed in 6 dense blocks and 2 transmit blocks (Fig. 2b indicating the architecture and data flow). The CNN took 16 resized 128-x128-pixel CT image sequences as input of each CT case, and generated a predicted pneumonia type with maximum probability as output across thousands of attached computing neurons. We defined the loss function as the weighted cross entropy between predicted probability and the true labels. Fine-tuned parameters of the network via back-propagation were optimized using batch size of 16, learning rate of 0.01, weight decay of 0.0001, momentum of 0.9, and epsilon of 0.00001. We conducted the training process utilizing a workstation equipped with 2 Tesla V100 GPUs, costing 6 hours to finish the task.

### Building AI model using federated learning

#### Data preparation

In experiment I, we trained with data collected from multiple centers of Tongji Hospital including Tongji Hospital Main Campus, Tongji Optical Valley Hospital, and Tongji Sino-French New City Hospital. We assigned each hospital to a federated client and place their local data on three different physical machines. In experiment II, besides data collected from above three hospitals, we added Wuhan Union Hospital as a new participant,

#### Federated model setup

For all experiments, we used the same architecture (3D-Densenet) with data-centralized training and the same set of local training hyperparameters for all clients with SGD optimizer: batch size of 35, learning rate of 0.01, momentum of 0.9 and weight decay of 5e-4. In experiment I, we set the number of federated rounds to 200 with one local epoch per federated round. A local epoch means each client train with its local data once before sending information to central server(cloud). We conducted the training process utilizing a workstation equipped with 3 Tesla V100 GPUs, costing 16 hours to finish. In experiment II, we set the number of federated rounds to 30 with one local epoch per federated round and start training with the global model coming from experiment I. For all experiments, we use the same evaluation metric with data-centralized training to check that our procedures are working properly. (In experiment II, we need to train 5 rounds before the model achieving the same performance with data-centralized training on test data from Wuhan Union Hospital).

#### Model aggregation

The server distributes a global model and receives synchronized weight updates (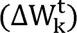) from all clients at each federated round. Due to each client train with one epoch per federated round, so we just average all the weight updates from the client with equal weight and update the global model.

#### Privacy-preserving setup

We use a variant of additively homomorphic encryption to achieve privacy-preserving, which called Learning with Errors (LWE)-based encryption. The encryption method allows us to leak no information of participants to the honest-but-curious parameter (cloud) server.

## Data Availability

All relevant data used for developing the initial model and federated models during the current study are not publicly available. We claimed that this study was approved by the Ethics Committee Tongji Hospital, Tongji Medical College of Huazhong University of Science and Technology to access this dataset for our research purpose.

http://www.ai-ct-covid.team/

## Data Availability

All relevant data used for developing the initial model and federated models during the current study are not publicly available.

## Model Availability

The online application of AI model is publicly available at http://www.ai-ct-covid.team/.

The initial model or offline APP is publicly available upon request at or through website http://www.ai-ct-covid.team/.

## Federated Learning Framework Availability

The source code can be accessed at https://github.com/HUST-EIC-AI-LAB/.

## Acknowledgements

This study was supported by HUST COVID-19 Rapid Response Call (No. 2020kfyXGYJ031, No. 2020kfyXGYJ093, No. 2020kfyXGYJ094) and the National Natural Science Foundation of China (61703171 and 81771801). This work was also supported in part by a grant from the National Cancer Institute, National Institutes of Health, U01CA242879, and Thammasat University Research fund under the NRCT, Contract No. 25/2561, for the project of “Digital platform for sustainable digital economy development”, based on the RUN Digital Cluster collaboration scheme.

## Author contributions

T.X., X.B., Z.L., and C.Z, conceived the work. Y.X., L.M., F.Y., K.M., J.Y., X.Y, C.S., Z.F., J.G., X.Z., R.H., C.Z., X. L., D.T., C.X., W.Z., D.Y., M.W., N.H., N.J.H., I.R.K., X.P., X.W.,J.B. designed and developed the models and analyses; Y.X., K.M., D.L.R., J.Z., and T.X. interpreted results; and K.M., J.W., P.M., D.L.R., J.Z., Z.L., and T.X. wrote the paper.

